# Cognition-oriented treatments and physical exercise on cognitive function in Huntington’s disease: Protocol for systematic review

**DOI:** 10.1101/2021.08.12.21261956

**Authors:** Katharine Huynh, Leila Nategh, Sharna Jamadar, Nellie Georgiou-Karistianis, Amit Lampit

## Abstract

**Introduction:** Cognitive impairments are prevalent in Huntington’s disease (HD), occurring many years prior to clinical diagnosis and are the most impactful on quality of life of patients. Cognitive interventions and exercise have been found to be efficacious in improving cognitive function in several clinical populations (e.g., older adults with mild cognitive impairment and dementia). However, the utility of cognitive interventions has not been systematically reviewed in HD. This systematic review aims to examine the efficacy of cognitive and physical interventions on cognitive function in HD.

**Methods:** Electronic databases (MEDLINE, EMBASE, PsycINFO, CENTRAL) were searched through till 10 May 2021 for interventional studies investigating the effect of cognition-oriented treatments and physical exercise on cognitive function in individuals with HD, compared to any control or no control. The primary outcome is change on objective measures of cognition. Additional outcomes include change in psychosocial, functional and neuroimaging measures. Variations of effects based on population and study factors will be considered. Risk of bias will be assessed using the Cochrane RoB 2 tool and ROBINS-I tool. Where appropriate, outcomes will be pooled using random-effects meta-analyses, heterogeneity will be examined using tau^2^ and I^2^ statistics, and moderators will be examined using meta-regression models.

**Discussion:** This review will systematically evaluate the efficacy of cognitive and physical interventions on improving cognitive function in HD. The eligibility criteria and planned analyses will allow for a comprehensive assessment of certainty in the evidence that will inform future trials and clinical practice.

**Registration:** This protocol was registered on PROSPERO (CRD42021259152).

## Introduction

### Rationale

Huntington’s disease (HD) is a genetically inherited neurodegenerative disease that causes progressive motor, psychiatric and cognitive decline [1,2]. Although diagnosis is based on motor symptoms, cognitive impairment is prevalent. In the pre-manifest phase of HD (prior to onset of diagnosable motor symptoms), mild cognitive impairment is reported in 40% of individuals [3], and this proportion increases to over 80% at the onset of manifest disease (where motor symptoms are sufficient for clinical diagnosis) [4]. Additionally, 5% of individuals are classified as having dementia at the stage of diagnosis [4]. Although cognitive symptoms are reported to have the greatest impact on quality of life beyond motor and psychiatric symptoms, there are no current pharmacological treatments available to improve or maintain cognition in HD [1,5].

Non-pharmacological interventions, specifically physical exercise and cognition-oriented treatments, have increasingly shown evidence of efficacy in improving cognition in other populations. These populations include healthy older adults [6-8], adults with mild cognitive impairment [6-9] and people with dementia [7,10,11]. Furthermore, some evidence suggests that physical exercise and cognitive interventions may affect cognition through different pathways, and multi-domain interventions such as physical exercise combined with cognitive training may offer greater benefits than single-domain interventions [12].

Existing reviews of the effects of non-pharmacological interventions such as physical exercise [13], physical therapy [14] and art-based rehabilitation [15,16] on cognitive function in HD have been largely inconclusive. This is partly attributed to reviews including only randomized controlled trials, leading to only a few studies being eligible for synthesis [13,15]. Further, the effect of cognitive interventions has been largely unexplored in HD. To our knowledge, only one systematic review of cognitive interventions in HD exists [17]. However, that review focused specifically on cognitive rehabilitation, was conducted almost ten years ago, and included only one study in HD. Finally, the effect of multi-domain interventions has not been assessed independently from single domain interventions [14]. A comprehensive review of the effect of cognitive, physical, and multi-domain interventions on cognitive function in HD, including randomized and non-randomized studies, is therefore warranted.

### Objectives

The primary aim of the review is to evaluate whether cognition-oriented treatments and physical exercise are associated with improved cognitive outcomes in individuals with HD, compared to any control or no control. The secondary aims are to evaluate the effects on psychosocial, functional and neuroimaging outcomes, and explore how these associations vary across different factors, such as participant and intervention characteristics.

## Materials and methods

This protocol adheres to the Preferred Reporting Items for Systematic Review and Meta-analysis Protocols (PRISMA-P) [18] and PRISMA 2020 guidelines [19]. The PRISMA-P 2015 checklist is provided as Supplementary file 1. The protocol was registered on PROSPERO on 15 July 2021 (registration number CRD42021259152). Important protocol amendments will be documented in the PROSPERO registry, and divergences from the protocol will be described in the final published review paper.

### Eligibility criteria

Studies fulfilling the following criteria are eligible.

#### Study design

Interventional studies including randomised controlled trials, quasi-experimental studies, pre-post studies with single group and interventional cohort studies will be considered. Case-control studies, cross-sectional studies and case studies will be excluded.

#### Participants

Studies including participants aged 18 or above, with clinically diagnosed manifest or pre-manifest HD are eligible. Studies that include other subgroups of participants are also eligible, but the review will only consider the subgroup with HD. Studies with participants aged below 18 will be excluded.

#### Interventions

Studies assessing physical exercise or cognition-oriented treatment, either alone, combined with each other, or with other non-pharmacological interventions, will be considered. The intervention must include a minimum of 3 hours of physical exercise or cognition-oriented treatment, either independently or combined with each other. Additionally, for interventions combining physical or cognitive interventions with other non-pharmacological interventions, the physical exercise and cognitive intervention components must constitute at least 50% of the total intervention. Physical exercise is defined as a structured activity that can improve endurance, strength, balance, or any combination of these aspects. This includes walking, cycling, resistance training, balance training, and mind-body exercises. Cognition-oriented treatments are defined as enhancing, restorative or compensatory interventions that aim to improve cognitive performance or reduce specific impairments. This includes cognitive stimulation, cognitive training, and cognitive rehabilitation [7]. Social prescribing interventions that do not provide an intervention will be excluded.

#### Comparators

Studies with any type of control group, including standard care, waitlist, no contact, or active control groups are eligible. Studies without control groups are also eligible. Control groups comprised of people without HD will be excluded.

#### Outcomes

Studies are eligible if they report change in objective measures of cognition (global or domain-specific) from baseline to post-intervention. Additional outcomes of interest include change in measures of psychosocial well-being (e.g., mood and quality of life), daily function (e.g., functional independence), and neuroimaging outcomes (e.g., brain volumes).

#### Other characteristics

No limits will be placed on year of publication, language, or country of publication.

### Information sources

Electronic databases (MEDLINE, EMBASE, PsycINFO, CENTRAL) were searched for eligible studies through 10 May 2021. Additional sources of grey literature included clinical trial registries via the WHO International Clinical Trials Registry Platform (searched on 4 June 2021), theses databases (ProQuest and EThOS; searched on 4 June 2021) and key conferences or meetings (Huntington Study Group, Huntington’s Disease Society of America, European Huntington’s Disease Network; searched on 6 June 2021). Reference lists of included articles and previous reviews will also be searched for additional articles. In instances where otherwise eligible studies are missing required outcome data, contact with authors will be attempted.

### Search strategy

The search strategy for MEDLINE is presented in Table 1. The search strategies for all databases, registries and websites are presented in Supplementary file 2. Search results were collated, and records were uploaded to an EndNote library. Duplicates were removed using EndNote and manual searching. Remaining records were uploaded to Covidence for screening.

**Table 1.**
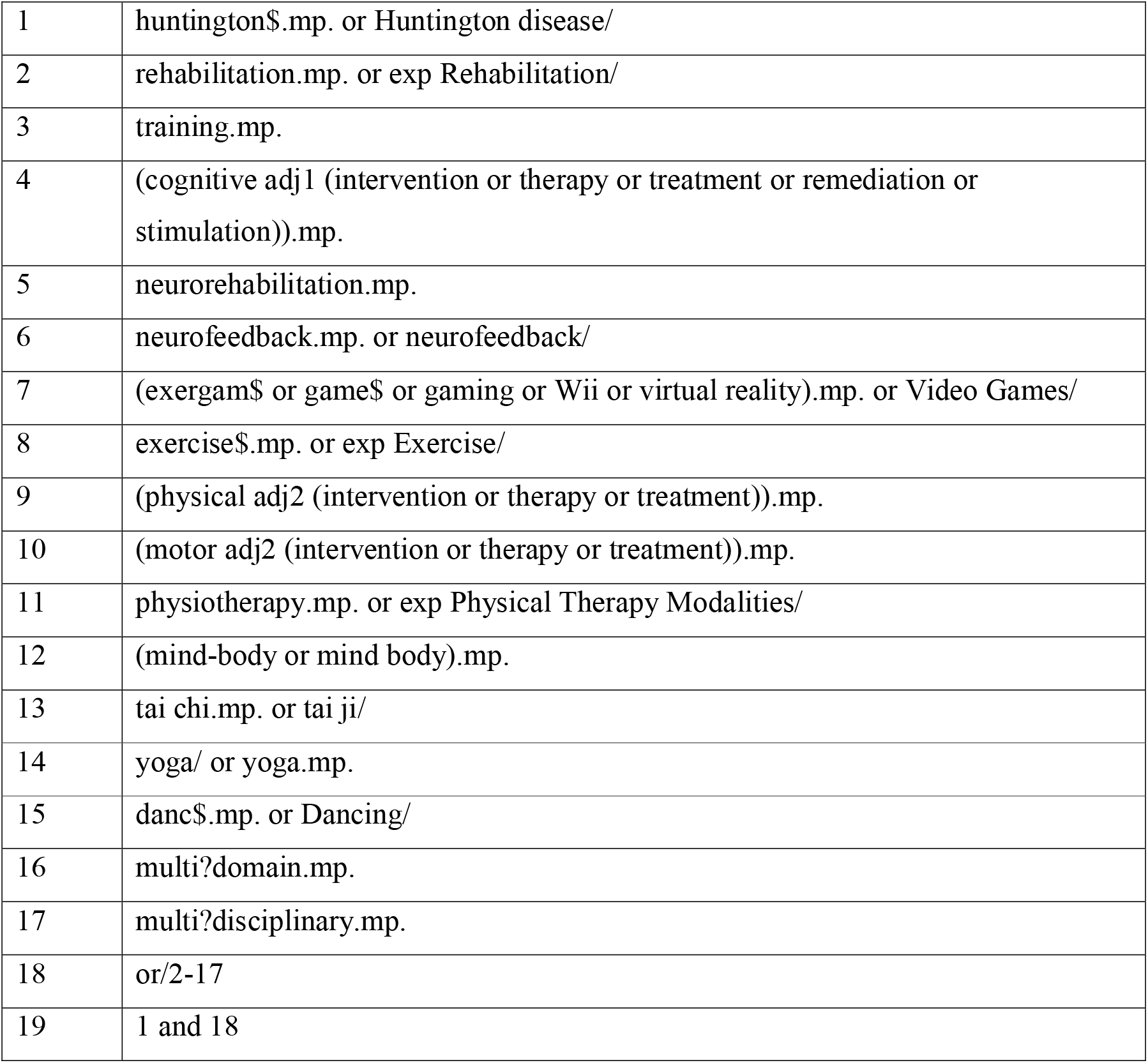
Search strategy for MEDLINE.

### Selection process

Screening based on title and abstracts, and full-text screening against eligibility criteria will be conducted independently by two reviewers. Disagreements at each stage will be resolved by consensus or a third reviewer.

### Data collection process

Data will be extracted to an Excel spreadsheet in duplicate by two reviewers. Disagreements will be resolved by consensus or resolved by a third reviewer. If data is missing or unclear, authors will be contacted to obtain data.

### Data items

#### Outcomes

The primary outcome is change in an objective measure of cognition (either global or domain-specific). Secondary outcomes include change in measures of psychosocial well-being (e.g., mood, depression, anxiety, quality of life), daily function (e.g., functional independence), and neuroimaging outcomes (e.g., total brain volume, caudate and putamen volumes, network connectivity).

Outcomes will be extracted for both experimental and control groups (if applicable). If multiple measures of an outcome are reported (e.g., multiple measures of working memory capacity), all eligible data will be extracted. If data is reported for multiple time points, we will extract data from pre-intervention to first post-intervention timepoints. If results from multiple analyses are available, data from intention-to-treat analyses will be preferred.

#### Other variables

The following information will also be extracted:

- Study information: Author, year of publication, study setting, study design
- Population: Mean age, percent male, mean CAG repeat length, mean disease burden score, disease stage (percent premanifest), disease duration (if applicable), mean United Huntington’s Disease Rating Scale total motor score
- Intervention: Type of intervention, intervention content, delivery format, supervision, session frequency (sessions/week), session length (minutes), total number of sessions, number of weeks, total duration of intervention (hours)
- Comparator (if applicable): Type of control, control activity
- Other information: Funding sources, conflicts of interest

If information is missing or unclear following attempts to contact authors, the study will be left out of relevant analyses (e.g., examining effects of specific study population or intervention characteristics).

### Study risk of bias assessment

Risk of bias will be assessed separately for each relevant outcome in each study using the Cochrane RoB 2 tool for randomised controlled trials [20] and ROBINS-I tool for non-randomised studies [21].

The RoB 2 tool considers:

1. Bias arising from the randomisation process
2. Bias due to deviations from intended interventions
3. Bias due to missing outcome data
4. Bias in measurement of the outcome
5. Bias in selection of the reported result
6. Overall bias

The ROBINS-I tool considers:

1. Bias due to confounding
2. Bias in selection of participants into the study
3. Bias in classification of interventions
4. Bias due to deviations from intended interventions
5. Bias due to missing data
6. Bias in measurement of outcomes
7. Bias in selection of the reported result
8. Overall bias

Risk of bias assessments will be conducted independently by two reviewers. Disagreements between the reviewers will be resolved by discussion, with involvement of a third review author where necessary. Study authors will be contacted if reports do not provide sufficient details to determine risk of bias.

Risk of bias information will be used when considering heterogeneity (if applicable) between study results, and in determining confidence in the body of evidence for each outcome.

### Effect measures

For each outcome, continuous data will be extracted as means and standard deviation. Dichotomous (events) data will be extracted as number of individuals who did and did not experience event in each group. If data is not in the desired format, data in other formats (e.g., effect sizes and confidence intervals) will be extracted and converted prior to synthesis of results.

### Synthesis methods

Studies will be grouped based on study design (randomised controlled trials separated from non-randomised studies), type of intervention and outcome. Studies with the same design, intervention and outcome will be synthesised together.

For continuous data, effects will be reported as standardised mean difference with 95% CI due to expected variability in the measures used across studies. For dichotomous data, effects will be reported as odds ratios or risk ratios.

Characteristics of all included studies will be presented in a summary table. Findings from syntheses will be reported in text, structured based on priority of outcomes (cognitive outcomes followed by additional outcomes), followed by type of intervention, and then study design (randomised controlled trials followed by non-randomised studies). Study results from each synthesis will be presented in summary tables, and forest plots will be included where meta-analysis is conducted.

Where at least 3 studies are available in a group (with the same design, type of intervention and outcome), results will be pooled using a multivariate random-effects meta-analysis to account for non-independence of effect sizes within studies [22,23]. Analyses will be conducted using the packages metafor, metaSEM, robumeta and clubSandwich for R.

Heterogeneity will be assessed using both the tau^2^ and the I^2^ statistics. If there is significant heterogeneity, we will use meta-regressions to explore heterogeneity in effect estimates according to risk of bias, study population and intervention characteristics [24].

Sensitivity analyses will be performed by removing studies at higher risk of bias and recalculating the pooled estimate, and additionally by comparing results from multilevel and robust variance estimation models.

If meta-analysis is not possible, findings will be synthesised narratively and similarly structured based on study design, type of intervention and outcome. Synthesis will similarly consider heterogeneity and causes of heterogeneity, including risk of bias, population and intervention characteristics.

### Reporting bias assessment

Evidence of small-study effect will be assessed by inspecting funnel plots of effect size versus standard error for each outcome [25]. Where there are at least 10 studies, we will use formal statistical methods to tests for small-study effects. If summary estimates are in the form of standardized mean differences, we will use Egger’s test [26]. If summary estimates are in the form of odds ratio, we will use the tests proposed by Peters, Sutton [27] and Rücker, Schwarzer [28] when there is low between-study heterogeneity (tau^2^ < 0.1) and substantial between-study heterogeneity (tau^2^ > 0.1), respectively. Where there are less than 10 studies, we will remove outliers and recalculate pooled effect sizes after their removal.

### Certainty assessment

Confidence in the body of evidence for each outcome will consider the risk of bias, heterogeneity, indirectness, imprecision, and evidence of small-study effects (including publication and reporting biases).

## Discussion

Cognitive impairments are prevalent in individuals with HD and greatly impact quality of life, but no effective pharmacological treatments are currently available [4,5]. This review will systematically evaluate the efficacy of cognition-oriented treatments and physical exercise on cognitive function in HD, as there is accumulating evidence of efficacy in other clinical populations. The eligibility criteria allow for the inclusion of randomized and non-randomized studies, and studies that include any control or no control, given the rarity of the disease and the small number of anticipated studies. The planned analyses will also examine the influence of risk of bias, study population, and intervention characteristics, to provide a comprehensive assessment of certainty in the evidence, and to inform future trials and clinical practice.

## Data Availability

Data sharing is not applicable to this manuscript as no new data were created or analyzed.

## Data availability statement

Data sharing is not applicable to this manuscript as no new data were created or analyzed.

## Contributions

KH led the design of the review and the drafting of the manuscript, LN edited the draft manuscript, SJ edited the draft manuscript, NGK contributed to the design of the review and edited the draft manuscript, AL contributed to the design of the review and edited the draft manuscript.

