## Supplementary file 2 for "Cognition-oriented treatments and physical exercise on cognitive function in Huntington’s disease: Protocol for systematic review"

**Supplementary file 2. Full search strategy for electronic databases, registries, and websites**

***Electronic databases***

Electronic databases (APA PsycInfo, EMBASE, CENTRAL, MEDLINE) were searched 10 May 2021 using the Ovid interface. Results were exported to an EndNote library and duplicates were removed. Remaining records have been uploaded to Covidence for screening by two independent reviewers. The search strategy for each database is presented below.

*APA PsycInfo*

| 1 | Huntingtons Disease/ or huntington$.mp. |
| --- | --- |
| 2 | exp Rehabilitation/ or rehabilitation.mp. |
| 3 | training.mp. |
| 4 | (cognitive adj1 (intervention or therapy or treatment or remediation or stimulation)).mp. |
| 5 | neurorehabilitation.mp. |
| 6 | neurofeedback.mp. or Neurotherapy/ |
| 7 | (exergam$ or game$ or gaming or Wii or virtual reality).mp. or exp Games/ |
| 8 | exp Exercise/ or exercise$.mp. |
| 9 | (physical adj2 (intervention or therapy or treatment)).mp. |
| 10 | (motor adj2 (intervention or therapy or treatment)).mp. |
| 11 | physiotherapy.mp. |
| 12 | (mind-body or mind body).mp. or Mind Body Therapy/ |
| 13 | tai chi.mp. |
| 14 | yoga.mp. |
| 15 | Dance Therapy/ or Dance/ or danc$.mp. |
| 16 | multi?domain.mp. |
| 17 | multi?disciplinary.mp. |
| 18 | or/2-17 |
| 19 | 1 and 18 |

*EMBASE*

| 1 | huntington$.mp. or Huntington chorea/ |
| --- | --- |
| 2 | rehabilitation.mp. or exp rehabilitation/ |
| 3 | exp training/ or training.mp. |
| 4 | (cognitive adj1 (intervention or therapy or treatment or remediation or stimulation)).mp. |
| 5 | neurorehabilitation.mp. |
| 6 | neurofeedback.mp. or neurofeedback/ |
| 7 | (exergam$ or game$ or gaming or Wii or virtual reality).mp. or video game/ |
| 8 | exercise$.mp. or exp exercise/ |
| 9 | (physical adj2 (intervention or therapy or treatment)).mp. |
| 10 | (motor adj2 (intervention or therapy or treatment)).mp. |
| 11 | physiotherapy.mp. or exp physiotherapy/ |
| 12 | (mind-body or mind body).mp. |
| 13 | tai chi.mp. or Tai Chi/ |
| 14 | exp yoga/ or yoga.mp. |
| 15 | danc$.mp. or dancing/ |
| 16 | multi?domain.mp. |
| 17 | multi?disciplinary.mp. |
| 18 | or/2-17 |
| 19 | 1 and 18 |

*CENTRAL*

| 1 | huntington$.mp. or Huntington disease/ |
| --- | --- |
| 2 | rehabilitation.mp. or exp Rehabilitation/ |
| 3 | training.mp. |
| 4 | (cognitive adj1 (intervention or therapy or treatment or remediation or stimulation)).mp. |
| 5 | neurorehabilitation.mp. |
| 6 | neurofeedback.mp. or neurofeedback/ |
| 7 | (exergam$ or game$ or gaming or Wii or virtual reality).mp. or Video Games/ |
| 8 | exercise$.mp. or exp Exercise/ |
| 9 | (physical adj2 (intervention or therapy or treatment)).mp. |
| 10 | (motor adj2 (intervention or therapy or treatment)).mp. |
| 11 | physiotherapy.mp. or exp Physical Therapy Modalities/ |
| 12 | (mind-body or mind body).mp. |
| 13 | tai chi.mp. or tai ji/ |
| 14 | yoga/ or yoga.mp. |
| 15 | danc$.mp. or Dancing/ |
| 16 | multi?domain.mp. |
| 17 | multi?disciplinary.mp. |
| 18 | or/2-17 |
| 19 | 1 and 18 |

*MEDLINE*

| 1 | huntington$.mp. or Huntington disease/ |
| --- | --- |
| 2 | rehabilitation.mp. or exp Rehabilitation/ |
| 3 | training.mp. |
| 4 | (cognitive adj1 (intervention or therapy or treatment or remediation or stimulation)).mp. |
| 5 | neurorehabilitation.mp. |
| 6 | neurofeedback.mp. or neurofeedback/ |
| 7 | (exergam$ or game$ or gaming or Wii or virtual reality).mp. or Video Games/ |
| 8 | exercise$.mp. or exp Exercise/ |
| 9 | (physical adj2 (intervention or therapy or treatment)).mp. |
| 10 | (motor adj2 (intervention or therapy or treatment)).mp. |
| 11 | physiotherapy.mp. or exp Physical Therapy Modalities/ |
| 12 | (mind-body or mind body).mp. |
| 13 | tai chi.mp. or tai ji/ |
| 14 | yoga/ or yoga.mp. |
| 15 | danc$.mp. or Dancing/ |
| 16 | multi?domain.mp. |
| 17 | multi?disciplinary.mp. |
| 18 | or/2-17 |
| 19 | 1 and 18 |

***Clinical trial registries and theses databases***

Clinical trial registries and thesis databases were searched on 4 June 2021. All results were reviewed by one reviewer to identify additional relevant studies that were not previously found in the electronic database search.

| **Registry/theses database** | **Search strategy** |
| --- | --- |
| WHO International Clinical Trials Registry Platform | Huntington |
| EThOS | Huntington |
| ProQuest | noft(Huntington*) AND (noft(rehabilitation OR (cognitive training) OR (cognitive intervention) OR (cognitive therapy) OR (cognitive treatment) OR (cognitive remediation) OR (cognitive stimulation) OR neurorehabilitation OR neurofeedback OR exergam* OR game* or gaming OR (virtual reality) OR exercise OR (physical intervention) OR (physical therapy) OR (physical treatment) OR physiotherapy OR mind-body OR (mind body) OR (tai chi) OR (tai ji) OR yoga OR danc* OR multi-domain OR multidomain OR multi-disciplinary OR multidisciplinary)) |

***Websites***

Websites of relevant organizations were searched on 6 June 2021. Each website was browsed for conference or meeting abstracts and presentations, affiliated publications and clinical trials. The pages (provided below) were scanned by one reviewer to identify additional relevant studies that were not previously found in the electronic database search.

| **Organisation name and URL** | **Pages searched** |
| --- | --- |
| Huntington Study Group  URL: https://huntingtonstudygroup.org | Past annual meetings  URL: https://huntingtonstudygroup.org/about/our-annual-meeting/past-annual-meetings/ |
|  | Current clinical trials  URL: https://huntingtonstudygroup.org/current-clinical-trials/ |
|  | Past clinical trials  URL: https://huntingtonstudygroup.org/past-clinical-trials/ |
| Huntington’s Disease Society of America  URL: https://hdsa.org/ | 2008-2020 annual conventions  URL: https://hdsa.org/about-hdsa/annual-convention/ |
|  | 2013-2020 research/investor reports  URL: https://hdsa.org/hd-research/research-investor-reports/ |
|  | Publications  URL: http://hdsa.org/shop/publications/ |
| European Huntington’s Disease Network  URL: http://www.ehdn.org | 2020-2021 plenary meetings  URL: http://www.ehdn.org/plenary-meeting/ |
|  | Research and publications  URL: http://www.ehdn.org/hd-clinicians-researchers/previous-ongoing-research/#Publication |
